# Oral ulcers of COVID-19 patients: a scoping review protocol

**DOI:** 10.1101/2021.01.22.21250326

**Authors:** Abanoub Riad, Julien Issa, Veronika Chuchmova, Simona Slezakova, Esraa Gomaa, Andrea Pokorna, Jitka Klugarova, Miloslav Klugar

## Abstract

**Objective:** This scoping review aims to systematically identify the types, characteristics, and possible pathophysiologic etiologies of the oral ulcers that emerge in COVID-19 patients.

**Introduction:** The oral cavity is a vulnerable niche for the most diverse microbial ecosystem in the human body; therefore, it presents a wide array of mucocutaneous complications that could indicate various acute and chronic conditions. The COVID-19-related oral conditions, including oral ulcers, had been widely debated as direct manifestations or indirect complications of the SARS-CoV-2 infection. According to a preliminary search of PROSPERO, MEDLINE, the Cochrane Database of Systematic Reviews and the *JBI Evidence Synthesis*, there is no published nor registered scoping review concerned with the oral ulcers of COVID-19 patients.

**Inclusion criteria:** The review will include studies included COVID-19 patients whose infection had been confirmed by RT-PCR testing regardless of infection severity and clinical course. Only the studies that reported COVID-19 patients with oral ulcers.

**Methods:** A three-phase search strategy will be carried out: an initial limited search, a full electronic search, and hand search using the reference lists of all included records. The main bibliographic databases of published literature will include MEDLINE® (Ovid), EMBASE (Elsevier), and Cochrane COVID-19 Study Register. All identified records will be managed using EndNote 9.2, and the titles and abstracts will be screened against the inclusion criteria before the full text of all potentially relevant studies will be examined. The data will be presented in tabular form, rating maps, and narrative summary.

**Registration:** This protocol had been pre-registered in Open Science Framework (OSF) Registries.^[1]^

## Introduction

The outbreak of the coronavirus disease (COVID-19) had required sensitive case definitions to be developed according to the dynamically growing epidemiologic evidence.^[2]^ The COVID-19-related extrapulmonary manifestations including the cutaneous lesions had been frequently reported since the beginning of this pandemic; however, it took several months until these lesions were recognized by the World Health Organization (WHO) within the clinical case definitions of COVID-19.^[2,3]^ According to the review of Daneshgaran et al. 2020, the vesicular rashes were suggested to be used for early diagnosis of COVID-19; while acral lesions were useful for epidemiological uses, and vascular rashes for the prognosis of the disease severity.^[4]^

The oral cavity is subjected to numerous environmental factors that make it vulnerable to mucocutaneous complications. This vulnerability is supported by the fact that the oral cavity is a locus of the most diverse microbial ecosystem in the human body; therefore, it can harbour accessible lesions that could be diagnostic for an array of systemic conditions.^[5]^ Ulceration of the oral epithelium, underlying connective tissues, or both may last for two weeks or more, and it can be triggered by chronic conditions including drug-induced oral ulcers, lichen planus, pemphigus vulgaris, mucous membrane pemphigoid, lupus erythematosus, Reiter’s syndrome, tuberculosis, eosinophilic ulcers, precancerous ulcers, and mycoses-related ulcers.^[6]^ Additionally, oral ulcers may be of acute nature and last for less than two weeks, e.g. traumatic ulcers, recurrent aphthous stomatitis (RAS), Bechet’s disease, allergic reactions, erythema multiform, necrotizing sialometaplasia, hematologic disorders-related ulcers, and bacterial and viral infections. The clinical presentation of some viral infections like herpes simplex virus, varicella-zoster virus, coxsackievirus, cytomegalovirus, Epstein-Barr virus, measles virus, and human immunodeficiency virus (HIV) includes typically oral ulcers and vesicles which may aid in their timely diagnosis and management.^[7]^

The oral symptoms of COVID-19 had been first reported in Lyon by Chaux-Bodard et al. 2020 as an irregular tongue ulcer which evolved concurrently with acral erythema preceding the COVID-19 respiratory symptoms thus suggesting a promising epidemiological and clinical significance of the oral lesions as inaugural symptoms.^[8]^ In the following weeks, hundreds of individual cases with different oral lesions were reported in multiple countries to explain the nature and the aetiology of these lesions, which were equally distributed in females vs males.^[9–11]^ The COVID-19-related oral symptoms could be depicted as drug reactions, viral enanthem, stress-induced lesions, bacterial or fungal co-infections, or inflammatory reaction.^[9,11]^ Heretofore, the pathophysiological hypotheses of COVID-19-related oral symptoms are supported by collateral observations that require further investigation; for instance, the hypothesis of viral enanthema is supported by the high expression of angiotensin-converting enzyme-2 (ACE-2) receptors in oral epithelium which serve as the primary entry of severe acute respiratory syndrome coronavirus-2 (SARS-CoV-2); also the inflammatory response hypothesis is supported by the increasing number of cases of the pediatric multisystem inflammatory syndrome temporally associated with SARS-CoV-2 (PMIS-TS).^[9,12]^ However, the onset of oral lesions had considerably varied across the reported cases due to lack of use of reference timepoints, it may indicate the variety of pathophysiologic etiologies and the natures of these lesions.^[9]^

The COVID-19-related oral ulcers had been reported in critically-ill,^[13,14]^ moderate,^[15]^ mild^[15–17]^ and asymptomatic cases^[16]^. Aphthous-like lesions were reported without necrosis and hemorrhagic crusts in younger patients, while in older patients with severe infection, the aphthous-like lesions had been necrotic and hemorrhagic.^[11]^ In a recent review of aphthous stomatitis in COVID-19 patients, the aphthous lesions were slightly predominant in females and people below 40 years old, and their onset ranged between 0 to 10 days since the emergence of the respiratory symptoms.^[15]^ The tongue was the most common location for ulcerative and erosive lesions of non-specific origin which appeared 3 days before the onset of respiratory symptoms in one of the cases.^[11]^ Herpetiform lesions were painful and emerged before, during, or after the respiratory symptoms.^[11]^

This scoping review’s primary objectives are to evaluate the breadth and depth of the evidence on oral lesions of COVID-19 patients and to evaluate their proposed pathophysiological courses and epidemiological and clinical significance. A preliminary search of PROSPERO, MEDLINE, the Cochrane Database of Systematic Reviews and the *JBI Evidence Synthesis* was conducted on January 20^th^, 2020, and no current or underway scoping reviews or systematic reviews on the topic were identified.

## Review question(s)

What are the possible etiologies, types, and characteristics of the oral ulcers that emerge in COVID-19 patients?

## Inclusion criteria

### Participants

This review will consider studies that include COVID-19 patients whose infection had been confirmed by means of RT-PCR.

### Concept

This review will consider studies that reported any type of oral ulcers for example: solitary vs. multiple, acute vs. recurrent, painful vs. painless, necrotic vs. non-necrotic, hemorrhagic vs. non-hemorrhagic, or contagious vs. non-contagious.

### Context

This review will consider studies that reported COVID-19 patients with mild, moderate, severe and critic clinical courses according to the Australian guidelines for clinical care of COVID-19 patients.^[18]^ The patients hospitalized either at intensive care or intermediate care (which provides more intensive monitoring and patient management than the general ward but less than is offered in the intensive care),^[19]^ and the patients who underwent home isolation will be included in this review.

### Types of sources

This scoping review will consider descriptive as well as analytical epidemiological studies. Furthermore, systematic reviews, narrative reviews and correspondence articles will be considered for inclusion in the proposed scoping review.

## Methods

The proposed scoping review will be conducted in accordance with the JBI methodology for scoping reviews.^[20]^

### Search strategy

The search strategy will locate both published and unpublished primary studies, reviews, and correspondence articles. A three-phase search strategy will be carried out. The first phase, which has been already completed, was an initial limited search of MEDLINE® (Ovid) and Google Scholar to identify relevant articles, followed by an analysis of the text words contained in the titles and abstracts of retrieved records, in addition to the index terms used to describe these records. A full search strategy for MEDLINE (PubMed) is stipulated in Appendix 1. Secondly, the search strategy will be developed according to the previous phase using all identified keywords and index terms, and it will be customized for each included information source. Finally, the bibliographical reference lists of all the included records will be screened for additional records.

Only studies published in English will be included. All the studies published since January 1^st^, 2020 will be included as the first cluster of COVID-19 cases was reported in Wuhan, China on December 30^th^, 2019.

## Information sources

The databases to be searched include:

- MEDLINE® (Ovid)

- EMBASE (Elsevier)

- Cochrane COVID-19 Study Register

### Study selection

Following the search, all identified records will be collated and uploaded into EndNote 9.2 (Clarivate Analytics, PA, USA) and duplicates removed. Following a pilot test, titles and abstracts will then be screened by two independent reviewers (A.R. and J.I.) against the inclusion criteria. The potentially relevant records will be retrieved in full and their citation details imported into the JBI System for the Unified Management, Assessment and Review of Information (JBI SUMARI; JBI, Adelaide, Australia).^[21]^ The full text of the selected records will be assessed in detail against the inclusion criteria by two independent reviewers (A.R. and J.I.). Reasons for exclusion of full-text records that do not meet the inclusion criteria will be recorded and reported in the scoping review. Any disagreements that arise between the reviewers at each stage of the selection process will be resolved through discussion or with a third reviewer (M.K.). The search results will be reported in full in the final scoping review and presented in a Preferred Reporting Items for Systematic Reviews and Meta-analyses for Scoping Reviews (PRISMA-ScR) flow diagram.^[22]^

### Data extraction

Data will be extracted from records included in the scoping review by two independent reviewers (A.R. and J.I.) using a draft charting form developed by the reviewers. The team will trial the charting form to ensure that all relevant results are extracted, and the table will be revise and amended when required during the data charting process. Modifications will be detailed in the full scoping review. Any disagreements that arise between the reviewers will be resolved through discussion or with a third reviewer. The primary studies’ authors will be contacted to request missing or additional data, where required. The data extracted will include specific details such as author(s), year of publication, country (where the cases were reported), study design, study population (age and gender), medical history, COVID-19 symptoms, ulcer characteristics (color, size, number, location, onset, and duration) and treatment. A draft extraction tool is provided (see Appendix 2).

### Data analysis and presentation

The extracted data will be presented in tabular form according to the objectives of this scoping review. Additionally, rating maps will be used to display the frequency of reported cases. The tabulated results will be accompanied by a narrative summary demonstrating how the results relate to the scoping review’s objectives.

## Data Availability

The data that support the findings of this study are openly available in Open Science Framework (OSF)

https://doi.org/10.17605/OSF.IO/JWAU4

## Ethical consideration

The ethical committee of Masaryk University – Faculty of Medicine has exempted this study from getting approval due to the fact that it uses open-access data.

## Data availability statement

The data that support the findings of this study are openly available in Open Science Framework at https://doi.org/10.17605/OSF.IO/JWAU4

## Appendix I: Search strategy

**MEDLINE (PubMed)**

Search conducted on February 6^th^, 2021, from (01.01.2020) to present.

| Search | Query | Field | Records retrieved |
| --- | --- | --- | --- |
| Limited to the date range from (01.01.2020) to present |  |  |  |
| #1 | COVID-19 OR SARS-CoV-2 OR 2019-nCoV OR severe acute respiratory syndrome OR coronavirus OR corona virus | All Fields | 100,753 |
| #2 | Oral OR mouth OR stomatitis OR cheilitis OR tongue OR lingual OR lip OR lip* OR labial OR buccal OR perioral OR palate OR palat* OR pharynx OR pharyn* OR oropharyn* OR gingiva OR tonsil OR lingual OR saliva* OR face OR facial OR salivary | All Fields | 109,532 |
| #3 | Ulcer* OR ulcer OR ulcerative OR aphthous OR aphthae OR herpes OR herpe* OR blister* OR blister OR erosion OR erosion* OR erosi* OR lesion OR lesion* OR vesicle OR vesicle* OR pressure ulcer OR injuries OR injury OR complication* OR complication OR manifestation OR manifestation* OR damage OR scare OR scare* OR sore OR sore* OR erythema multiforme OR lichen planus OR lichen* OR pemphigus OR pemphigoid OR erythem* OR varicella OR coxsackie OR cytomegalovirus OR Epstein–Barr | Title / Abstract | 257,772 |
| #4 | #1 AND #2 AND #3 |  | 1,038 |

## Appendix II: Data extraction instrument

| Author, Date, Location | Number of patients | Gender | Age | Medical anamnesis | COVID-19 symptoms | COVID-19 treatment | Ulcer characteristics |  |  |  | Ulcer Treatment |
| --- | --- | --- | --- | --- | --- | --- | --- | --- | --- | --- | --- |
|  |  |  |  |  |  |  | Location | Number / Size | Necrosis / Hemorrhage | Onset / Duration |  |

